# Serotypes and Serogroups implicated in Bacterial meningitis across Ghana

**DOI:** 10.1101/2022.06.10.22276133

**Authors:** Enoch Weikem Weyori, Abass Abdul-Karim, John Bertson Eleeza, Nkrumah Bernard, Braimah Baba Abubakari, Hilarius Asiwome Kosi Abiwu, Kuugbee Dogkotenge Eugene, Adadow Yidaana, Benjamin Nuertey, Adatsi Richard Kojo, Stebleson Azure, Sylvester Mensah, Etowi Boye Yakubu, Valentine Koyiri Cheba, Alentis Aba-ifaa Dombadoh

## Abstract

**Background:** Bacterial meningitis is a dangerous infection that can kill children and adults alike. An estimated 1.2 million instances of bacterial meningitis are anticipated to occur globally each year, despite the discovery of polysaccharide and conjugate vaccines in recent years [1]. The study aimed to find out which bacteria isolates are linked to meningitis and to compare the culture method to the rT-PCR method.

**Method:** This study examined data on bacterial meningitis from the Ghana Health Service’s Public Health Division from 2015 to 2019. Patients’ information was collected from case-based forms held at the Tamale Public Health and Reference Laboratory. The data from the case-based forms was transcribed into a pre-designed Microsoft Excel template. For analysis, the data was cleaned and loaded into Minitab version 18.

**Results:** There were 2,446 CSM cases documented in all, 34.4 percent were confirmed. Males (52.7%) were more suspected than females (47.3%). Age group 15-44 were affected most (37.5%). The predominant pathogens were *Neisseria meningitidis* (W135) and *Streptococcus pneumoniae* (St.1) with NMX, St.14, St.19, and St. 12F/12A/12b as new emerging strains.

Notwithstanding that, low NPV (72.6%), NLR (0.69), and sensitivity (31.8%) of culture method for detected pathogens of bacterial meningitis were found to produce a statistically significant false negatives compared to the gold standard (rT-PCR).

**Conclusion:** Emergence of new strains of bacterial meningitis and the false negative results chained out by the culture method is alarming.

## Introduction

Meningitis is a devastating disease with a high case fatality rate and leading to serious long-term **c**omplications. It causes up to 20% of deaths in victims and also accounts for extreme neurological sequelae (influencing 12-15% of survivors) in non-industrial nations. Even though several bacteria might cause the sickness, three isolates (*Streptococcus pneumoniae* (Sp), *Neisseria meningitidis* (Nm), and *Haemophilus influenzae* type b (Hib)) contribute to a significant proportion accounting for more than one million mortalities every year, principally among kids and adults (WHO, 2002). In the countries in the ‘African meningitis belt,’ from Ethiopia to Senegal, where incidence rates of the disease are significantly greater, an epidemic of bacterial meningitis disease is a major public health hazard [2]. Meningococcal infections, responsible for severe annual outbreaks of meningitis, are the most feared, with an incidence rate as high as 1000 cases per 100 000 population unlike *Streptococcus pneumoniae* that contributes approximate 17 cases per 100,000 population annually with a CFR exceeding 73% in children under five years. Though *H. influenzae* meningitis is rare in populations of adolescents and adults, rates of meningitis due to Hib are highest in children less than five years of age, with an estimated incidence rate of 31 cases per 100,000 accounting for higher mortality rates than found in *Neisseria meningitis* [2]–[5][2]–[5].

Notwithstanding that, an additional concern is related to the recent change in the epidemiological pattern of meningococcal outbreaks due to the emergence of serogroup W135 as an epidemic strain as well as the serotypes of *Streptococcus pneumoniae* in populations of developing countries due to the enhancement of the PCR assay technique [6]. The examination of bacterial meningitis relies upon the blood and CSF tests conducted by rapid RDT test, culture and PCR methods. Observational anti-microbial treatment is to be started quickly founded on the clinical discoveries and rapid test results [7]. For a successful treatment of bacterial meningitis, the microorganisms and their anti-toxin weakness examples ought to be quickly identified [8]. While the CSF culture is the highest quality level in the finding of bacterial meningitis, the low bacterial development rates especially in the patients who have gotten anti-microbial treatment before the lumbar cut (LP) required the advancement of new test methods [9]. Nucleic corrosive enhancement tests, for example, the PCR can distinguish limited quantities of microbe DNA freely from the development of the microorganism causing the disease [10].

In the light of these revelations, implementation of the PCR assay for diagnosis and surveillance of bacterial meningitis in Ghana [11]–[13], Niger [14], [15] and Burkina Faso [16], among the most stricken countries, is an important milestone for the meningitis national control programmes. This paper reports on the bacterial isolates implicated in the meningitis as well as benefits of the PCR assay compared with culture diagnostic technique across the meningitis belt in Ghana.

### Study Design

The research constitutes a retrospective institutional cross-sectional study conducted within the northern zone of Ghana taking into account data from 2015 to 2019.

### Study Setting

The source of data for the study included the Zonal Public Health Laboratory, Tamale (ZPHL) under the Ghana Health Service, Northern regional directorate. ZPHL is a reference laboratory for bacterial meningitis in Ghana. It also serves as the reference public health laboratory for the northern zone with respect to diseases of Public Health importance.

### Study Population

The study population was made up of all suspected, probable, and confirmed cases of bacterial meningitis. All cases from Upper East, Upper West, Savannah, North-East, and Northern regions were part of the geographical composition of the study.

### Clinical case definition

An illness with sudden onset of Fever (>38.5°C rectal or >38.0°C axillary) with one or more of the following symptoms: neck stiffness, altered consciousness, additional meningeal sign, or petechial or purpureal rash (GHS, 2019). In patients less than one (1) year, suspect meningitis when fever accompanied by bulging fontanelle (GHS, 2019).

### Laboratory criteria for diagnosis

Within 72 hours of clinical manifestation, positive CSF antigen detection or positive culture at the district or facility level was validated by PCR test (GHS, 2019).

#### Case classification

Suspected case is one that meets the clinical case definition; probable case is one that meets the clinical case definition and has turbid CSF (with or without positive Gram stain) or is part of an ongoing epidemic with an epidemiological link to a confirmed case; and confirmed case is one that has laboratory confirmation.

### Sample Size determination

From 2015 to 2019, all probable instances of bacterial meningitis were included in the study, hence no sample size was calculated.

### Data Analysis

Data was retrieved from the case surveillance forms and entered into a Microsoft Excel spreadsheet. For analysis, the data was cleaned, validated, and exported to Minitab version 18. A descriptive analysis was carried out and the results were reported in frequency tables. Statistical significance testing was done using the paired samples sign test with the sensitivity and specificity of culture method determined; positive predictive value (PPV) and negative predictive value (NPV) were calculated. Likelihood ratios for PPV and NPV were also estimated to determine the accuracy of the culture method.

### Inclusion Criteria

The study included all patients who met the case definition criteria for bacterial meningitis as determined by the Disease Control and Surveillance Unit of the Ghana Health Service

### Exclusion Criteria

All patients with inadequately filled case investigation forms, cases that were not having samples accompanying the case investigation forms and cases that had no culture and PCR results.

### Patient and Public Involvement

The study had no patient or public participation because it was a retrospective study.

## Findings and Results

Age 15 – 44 had majority of suspect and confirmed cases across the trend years; males had greater proportion for suspected (1288; 52.7%) and confirmed (466; 36.2%) cases compared to females with 47.3% of suspected cases and 32.5% of confirmed cases. Most suspected cases were from northern (993; 40.6%) and upper west (993; 40.6%) regions with majority confirmed cases from northern region (427; 43.0%) (Table 1).

**Table 1:** Demographic Characteristics of Cases

| Variable | Level | Negative<br>(n=1604) | Positive<br>(n=842) | Total<br>(n=2446) |
| --- | --- | --- | --- | --- |
| Age Category | < 5 | 316 (71.5) | 126 (28.5) | 442 (18.1) |
|  | 5-14 | 358 (53.2) | 315 (46.8) | 673 (27.5) |
|  | 15-44 | 665 (67.8) | 316 (32.2) | 981 (40.1) |
|  | 45-59 | 136 (74.7) | 46 (25.3) | 182 (7.4) |
|  | 60 + | 129 (76.8) | 39 (23.2) | 168 (6.9) |
| **Sex** | Male | 822 (63.8) | 466 (36.2) | 1288 (52.7) |
|  | Female | 782 (67.5) | 376 (32.5) | 1158 (47.3) |
| **Regions** | Northern | 566 (57.0) | 427 (43.0) | 993 (40.6) |
|  | Upper East | 283 (61.5) | 177 (38.5) | 460 (18.8) |
|  | Upper West | 755 (76.0) | 238 (24.0) | 993 (40.6) |

### Sex and age distribution

Table 2 indicates majority (466; 55.3%) were males with only 2015 (30; 51.7%) having higher counts for females. Nevertheless, the age categorization saw age 15-14 years with the highest positive cases in 2015 (37.9%) to 2017 (37.2%) compared to 15-44 years having the highest positivity rate in 2018 (38.4%) to 2019 (44.4%).

**Table 2:**
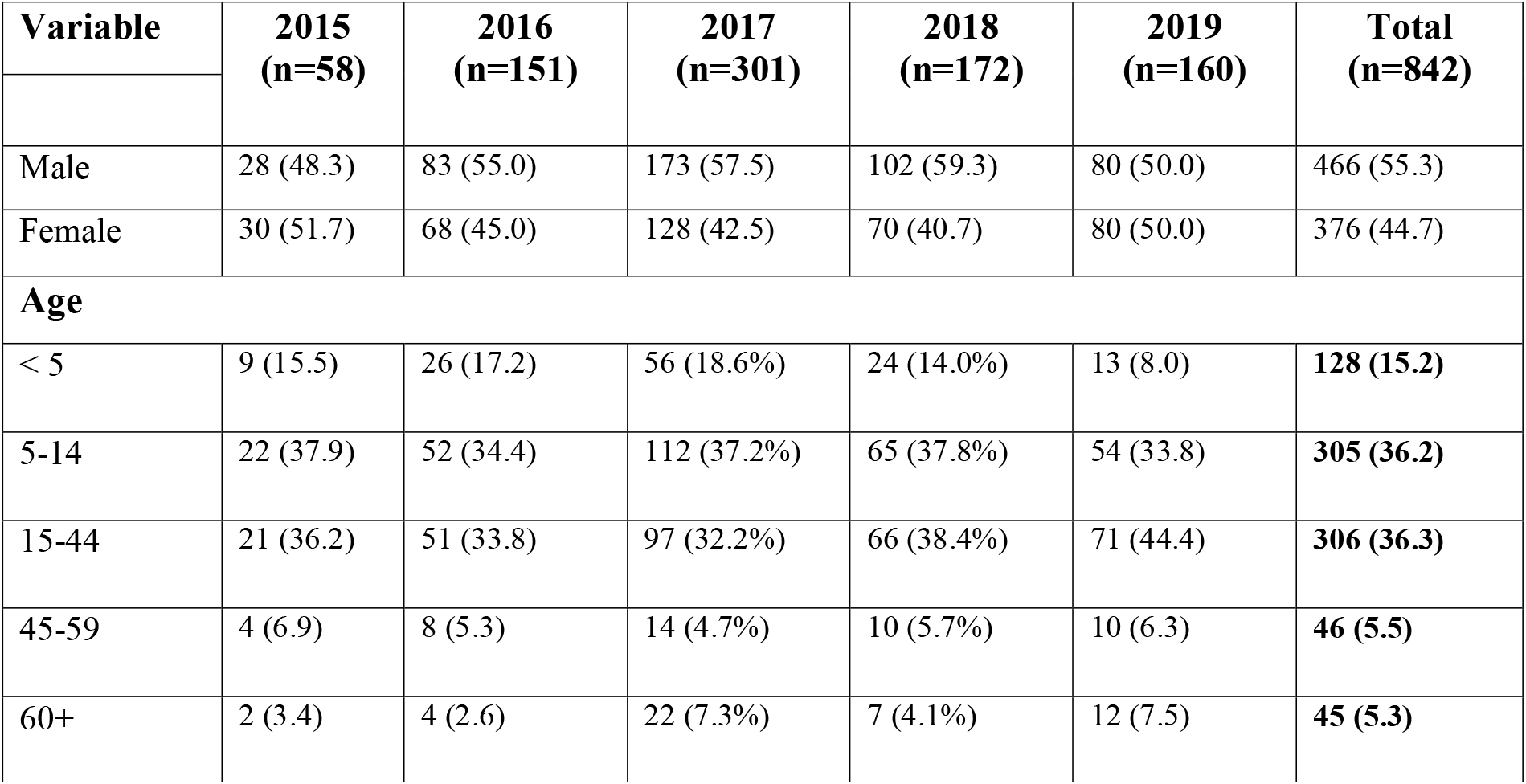
Sex and age distribution of confirmed cases across the years

| **Variable** | **2015**<br>**(n=58)** | **2016**<br>**(n=151)** | **2017**<br>**(n=301)** | **2018**<br>**(n=172)** | **2019**<br>**(n=160)** | **Total**<br>**(n=842)** |
| --- | --- | --- | --- | --- | --- | --- |
| Male | 28 (48.3) | 83 (55.0) | 173 (57.5) | 102 (59.3) | 80 (50.0) | 466 (55.3) |
| Female | 30 (51.7) | 68 (45.0) | 128 (42.5) | 70 (40.7) | 80 (50.0) | 376 (44.7) |
| **Age** |  |  |  |  |  |  |
| < 5 | 9 (15.5) | 26 (17.2) | 56 (18.6%) | 24 (14.0%) | 13 (8.0) | **128 (15.2)** |
| 5-14 | 22 (37.9) | 52 (34.4) | 112 (37.2%) | 65 (37.8%) | 54 (33.8) | **305 (36.2)** |
| 15-44 | 21 (36.2) | 51 (33.8) | 97 (32.2%) | 66 (38.4%) | 71 (44.4) | **306 (36.3)** |
| 45-59 | 4 (6.9) | 8 (5.3) | 14 (4.7%) | 10 (5.7%) | 10 (6.3) | **46 (5.5)** |
| 60+ | 2 (3.4) | 4 (2.6) | 22 (7.3%) | 7 (4.1%) | 12 (7.5) | **45 (5.3)** |

### Isolates implicated in Bacterial meningitis

Table 3 shows the real time Rt-PCR confirmed bacterial causative agents of meningitis over the years studied. It was denoted that, from the 842 cases, *Streptococcus Pneumonia (Spn)* accounted for 53.0% (446) of all confirmed cases of bacterial meningitis trailed by *Neisseria meningitis (Nm)* (43.7%; 368) and *Haemophilus influenzae* (3.3%; 28) from 2015 to 2019. Except for 2015 and 2016 where *Neisseria meningitis* was equal and more as causative agent of bacterial meningitis, *Streptococcus pneumonia* was implicated in most of the cases with *Haemophilus influenzae* accounting for a small proportion of the cases.

**Table 3:** Rt-PCR pathogens detected over the trend years

| **Rt-PCR Results** | **2015**<br>**(n=58)** | **2016**<br>**(n=151)** | **2017**<br>**(n=301)** | **2018**<br>**(n=172)** | **2019**<br>**(n=160)** | **Total**<br>**(n=842)** |
| --- | --- | --- | --- | --- | --- | --- |
| NM | 28 (48.3) | 97 (64.2) | 125 (41.5) | 62 (36.0) | 56 (35.0) | **368 (43.7)** |
| SPN | 28 (48.3) | 51 (33.8) | 166 (55.1) | 102 (59.3) | 99 (61.9) | **446 (53.0)** |
| HI | 2 (3.4) | 3 (2.0) | 10 (3.3) | 8 (4.7) | 5 (3.1) | **28 (3.3)** |

NM: *Neisseria meningitis*, SPN: *Streptococcus pneumoniae*, HI: *Haemophilus influenzae*

### Serogroups of Neisseria meningitis

Table 4 indicates the cross tabulation of the serogroups of *Neisseria meningitis* causing bacteria meningitis from 2015 to 2019. Of the 368 cases of Neisseria meningitis, serogroup NmW accounted for about 84.0% (309) of the cases followed by serogroups NmX (11.7%; 43), NmNG (2.7%; 10), NmC (1.1%; 4) and NmB (0.5%; 2)

**Table 4:** Rt-PCR Results of Neisseria meningitis Serogroups

| Isolates (NM) | Years |  |  |  |  | Total<br>(n=368) |
| --- | --- | --- | --- | --- | --- | --- |
|  | 2015<br>(n=28) | 2016<br>(n=97) | 2017<br>(n=125) | 2018<br>(n=62) | 2019<br>(n=56) |  |
| NmB | 0 (0.0) | 0 (0.0) | 1 (50.0) | 1 (50.0) | 0 (0.0) | 2 (0.5) |
| NmC | 0 (0.0) | 1 (25.0) | 3 (75.0) | 0 (0.0) | 0 (0.0) | 4 (1.1) |
| NmW | 28 (9.1) | 95 (30.7) | 119 (38.5) | 39 (12.6) | 28 (9.1) | 309 (84.0) |
| NmX | 0 (0.0) | 1 (2.3) | 2 (4.7) | 17 (39.5) | 23 (53.5) | 43 (11.7) |
| NG | 0 (0.0) | 0 (0.0) | 0 (0.0) | 5 (50.0) | 5 (50.0) | 10 (2.7) |

Notwithstanding that, indications shows that 2017 had the highest frequency of *Neisseria meningitis* cases with 125 representing 34.0 percent whiles 2015 recorded the least frequency of cases with 28 (7.6%).

### Serotypes of Streptococcus pneumoniae (2015-2019)

Table 5 relates to the cross tabulation of polymerase chain reaction results for serotypes of *Streptococcus Pneumoniae* causes of meningitis over the five years period under review. In indication, *Spn Serotype 1 (St. 1)* recorded the majority of cases representing almost half of the cases (49.6%). Furthermore, relative to *Spn* cases recorded over the period, it was realized that *St 18C/18B/18A/18F* had the least counts of 1 (0.2%) case. Other cases like *St. 12F/12A/12B/44/4* and *St. 14* had frequencies of 45 (10.1%) and 21(4.7%) respectively. It was worth noting that non-typable cases of *Streptococcus pneumoniae* cases had a significant frequency of 61 (13.7%).

**Table 5:** Rt-PCR Results serotypes against the trend years

| Rt-PCR Results | Years |  |  |  |  | Total<br>(446) |
| --- | --- | --- | --- | --- | --- | --- |
|  | 2015<br>(n=28) | 2016<br>(n=51) | 2017<br>(n=166) | 2018<br>(n=102) | 2019<br>(n=99) |  |
| St. 1 | 17 (7.7) | 31 (14.0) | 87 (39.4) | 56 (25.3) | 30 (13.6) | 221 (49.6) |
| St. 11A/11D | 0 (0.0) | 0 (0.0) | 0 (0.0) | 2 (40.0) | 3 (60.0) | 5 (1.1) |
| St. 12F/12A | 0 (0.0) | 0 (0.0) | 2 (50.0) | 2 (50.0) | 0 (0.0) | 4 (0.9) |
| St. 12F/12A/12B | 0 (0.0) | 0 (0.0) | 0 (0.0) | 2 (14.3) | 12 (85.7) | 14 (3.1) |
| St. 12F/12A/12B/44/4 | 0 (0.0) | 10 (22.2) | 21 (46.7) | 6 (13.3) | 8 (17.8) | 45 (10.1) |
| St. 14 | 1 (4.8) | 0 (0.0) | 4 (19.0) | 2 (9.5) | 14 (66.7) | 21 (4.7) |
| St. 15A/15F | 0 (0.0) | 0 (0.0) | 2 (100.0) | 0 (0.0) | 0 (0.0) | 2 (0.4) |
| St. 18C/18B/18A/18F | 0 (0.0) | 0 (0.0) | 1 (100.0) | 0 (0.0) | 0 (0.0) | 1 (0.2) |
| St. 19A | 0 (0.0) | 0 (0.0) | 2 (16.7) | 0 (0.0) | 10 (83.3) | 12 (2.7) |
| St. 19F | 0 (0.0) | 0 (0.0) | 1 (100.0) | 0 (0.0) | 0 (0.0) | 1 (0.2) |
| St. 23F | 1 (5.3) | 0 (0.0) | 18 (94.7) | 0 (0.0) | 0 (0.0) | 19 (4.3) |
| St. 3 | 1 (5.9) | 0 (0.0) | 8 (47.1) | 3 (17.6) | 5 (29.4) | 17 (3.8) |
| St. 33F/33A/37 | 0 (0.0) | 0 (0.0) | 3 (50.0) | 1 (16.7) | 2 (33.3) | 6 (1.3) |
| St. 4 | 0 (0.0) | 0 (0.0) | 1 (50.0) | 1 (50.0) | 0 (0.0) | 2 (0.4) |
| St. 5 | 8 (61.5) | 1 (7.7) | 4 (30.8) | 0 (0.0) | 0 (0.0) | 13 (2.9) |
| St. 6A/6B | 0 (0.0) | 0 (0.0) | 0 (0.0) | 2 (100.0) | 0 (0.0) | 2 (0.4) |
| NT | 0 (0.0) | 9 (14.8) | 12 (19.7) | 25 (41.0) | 15 (24.5) | 61 (13.7) |

Notwithstanding, the year that recorded the highest frequency of *Spn* cases was 2017 with 166 (37.2%) whiles 2015 recorded the least with 28 (6.3%).

### Comparison of PCR and Culture results Outcome for Equality

The hypothesis was therefore developed as;

**H**_**0**_: There is no statistically significant difference between the culture test results and that of the PCR test results

**H**_**1**_: There is a statistically significant difference between the culture test results and that of the PCR test results

A total of 2446 patients were examined using the two methods, *pathogens (11*.*1%) were* both isolated in the CSF culture and detected in the rT-PCR method. Twenty-four percent were detected by rT-PCR method but not detected by the CSF culture method. There was 0.2% of the samples in which the CSF culture was positive although the rT-PCR was negative (Table 6).

**Table 6:** Comparison of the CSF culture and the Rt-PCR results.

| Method |  |  | Rt-PCR test |  | PPV |
| --- | --- | --- | --- | --- | --- |
|  |  |  | Positive | Negative |  |
| CSF culture | Positive | Count | 272 | 5 | 277 |
|  |  | % of N | 11.1 | 0.2 | 65.6 |
|  | Negative | Count | 570 | 1599 | 842 |
|  |  | % of N | 23.3 | 65.3 | 34.4 |
| Total |  | Count | 2169 | 277 | 2446 |
|  |  | % of Total | 88.7 | 11.3 | 100.0 |

Table 7 represents the sensitivity and specificity of the CSF culture test method. In order to assess the performance of CSF culture, results were compared with Rt-PCR as a gold standard. Accordingly, the diagnostic accuracy of CSF culture including sensitivity (31.8%), specificity (98.2%), positive predictive value (90.5%), and negative predictive value (72.6%). Notably, the positive predictive likelihood ratio of CSF culture was 17.7 whiles the negative likelihood ratio was 0.69.

**Table 7:**
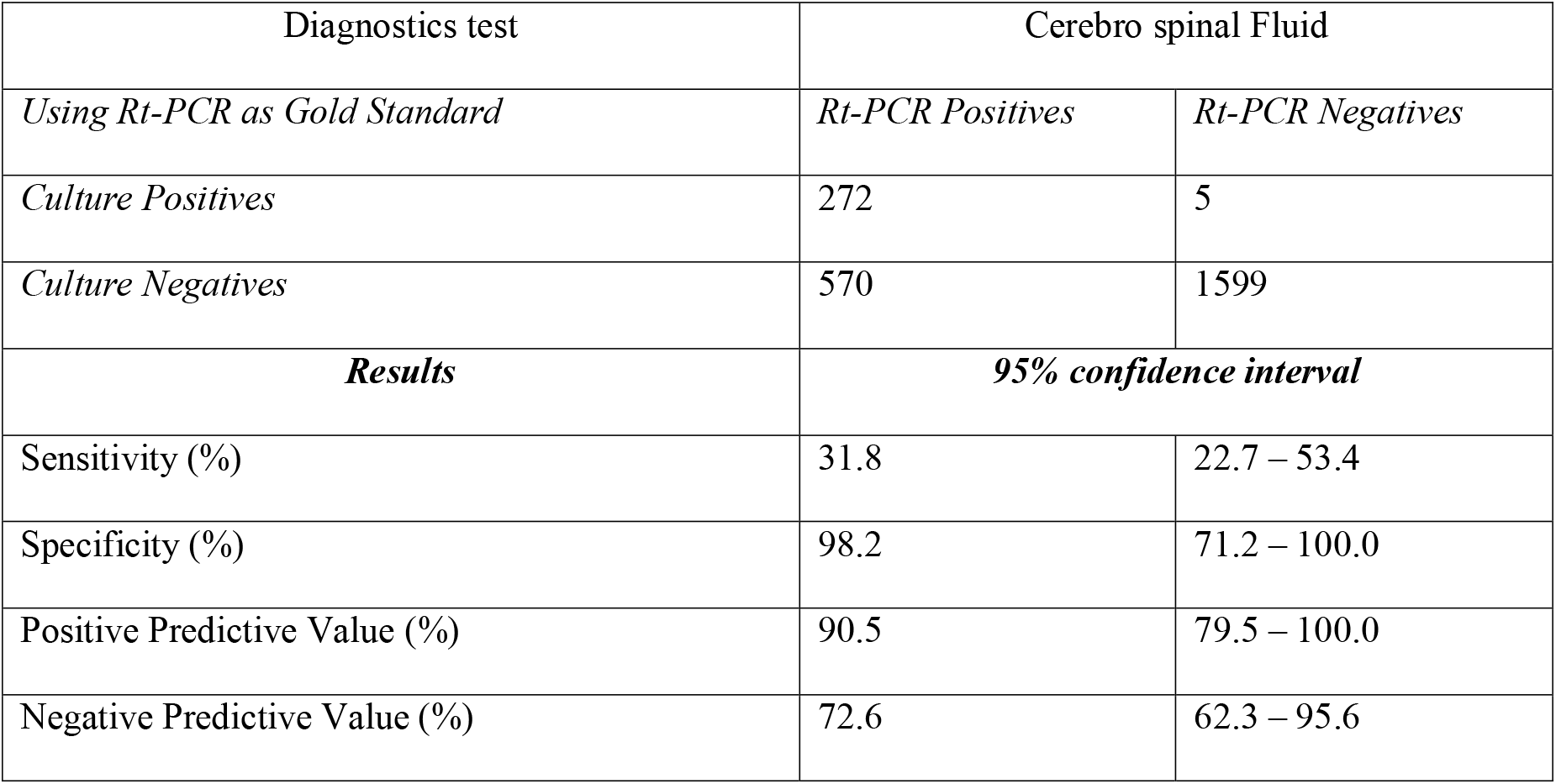
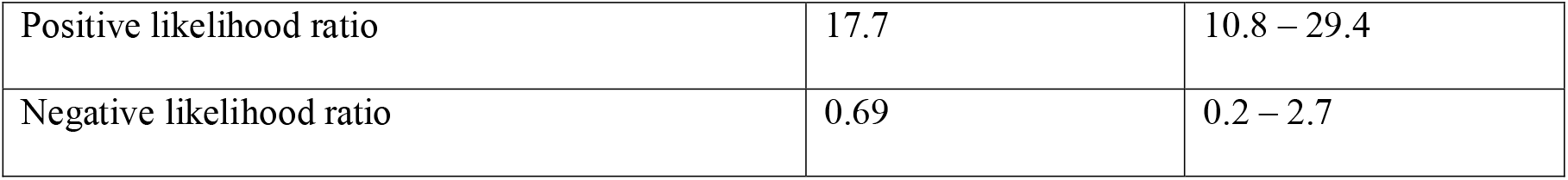
Diagnostic accuracy comparing PCR and culture for CSF samples

| Diagnostics test | Cerebro spinal Fluid |  |
| --- | --- | --- |
| *Using Rt-PCR as Gold Standard* | *Rt-PCR Positives* | *Rt-PCR Negatives* |
| *Culture Positives* | 272 | 5 |
| *Culture Negatives* | 570 | 1599 |
| **Results** | **95% confidence interval** |  |
| Sensitivity (%) | 31.8 | 22.7 – 53.4 |
| Specificity (%) | 98.2 | 71.2 – 100.0 |
| Positive Predictive Value (%) | 90.5 | 79.5 – 100.0 |
| Negative Predictive Value (%) | 72.6 | 62.3 – 95.6 |
| Positive likelihood ratio | 17.7 | 10.8 – 29.4 |
| Negative likelihood ratio | 0.69 | 0.2 – 2.7 |

**Table 8:** Shows Paired Sample Sign Test for difference in test outcomes

| Method Verification | Measurement | Frequencies<br>(n = 2446) | Percent<br>(%) |
| --- | --- | --- | --- |
| Culture results for Signed Values - PCR results for Signed Values | Negative Differences <sup>a</sup> | 570 | 23.3 |
|  | Positive Differences <sup>b</sup> | 5 | 0.2 |
|  | Ties <sup>c</sup> | 1871 | 76.5 |
|  | Z – value | -22.596 |  |
|  | Asymptomatic. Sig. (2-tailed) | .000 |  |

Majority (1823; 74.5%) of results remain symmetric whiles few (570; 23.3%) are parallel for rT-PCR and culture results. Also, 0.2% of positive culture results tested negative for rT-PCR with a significant p-value (p<0.00). This suggests rejection of H_0_ indicative of a statistically significant difference between results chained out by the Culture and rT-PCR methods.

## Discussion of Results

The number of confirmed cases were a 34.4% of the total cases suspected over the period. *Neisseria meningitidis* (43.7%) and *Streptococcus pneumonia* (53.0%) were the most predominant pathogens or isolates associated with bacterial meningitis in the three northern regions contributing approximately 95% of confirmed cases. In addition, the pathogens of *Haemophilus influenzae* were 3.3%. Across the trend years, notably were *Streptococcus pneumoniae* with highest positivity rates whereas *Haemophilus influenzae* had the lowest positivity rates. This similarity of pathogens across the trend years could be as a result of the introduction of vaccines to control some strains of the pathogens (*Neisseria meningitis* W135). These findings are in well match with CDC, (2012) which stated the most common strains of bacterial meningitis seen in West Africa over the past decades are basically *Streptococcus pneumoniae, Neisseria meningitidis, Haemophilus influenzae*, and *Group B Streptococcus* [17].

Notwithstanding that, *Neisseria meningitidis* had common strains within the five-year period to be B (0.5%), C (1.1%), W135 (84.0%), and X (11.7%). It was also worth noting that the most common strain of this group is the *W135*, whereas *NmX* is identified as the new strain springing up in the meningitis belt of Ghana. Some strains of *Neisseria meningitidis* that could not be grouped by the laboratory due to limited capacity for testing. The results assent to Nyarko who uncovered that *Neisseria meningitides W135* serogroup which was once rare in Ghana, with only four cases reported in 2004 has now become a common cause of outbreaks in the region [18]. These results also relate and conforms to that of CDC, (2012) which stated that the most prevalent and common strains of *Neisseria meningitidis* in Africa is the serogroup A, B, C, W135, X, and Y. Contrary to previous findings that uncovered 80-85% of all of *Neisseria meningitis* in the African Meningitis belt are due to serogroup A (WHO, 2015), a different trend is observed in this study where *Neisseria meningitis W 135* constituted up to 84.0% of *Nm* isolates.

*Streptococcus pneumoniae* in recent times have experienced spikes in number of cases and accounted for 53.0% of the total confirmed cases across the five-year period under review with serotype 1 (St. 1) accounting for majority (49.6%) cases. Notwithstanding that, some of the strains (Serotypes) of *Streptococcus pneumoniae* (13.7%) were non-typable. The evidence implies that approximately 70.0% of *Streptococcus pneumoniae* are accounted for by the most common serotypes (St.1, St.12F/12A, St.12F/12A/12B/44/4, and St.14). The emergence of new strains is evident in St 12F/12A/12B, St 14, and St 19. This finding of this study agrees with other researchers who observed a similar increase in incidence of *Streptococcus pneumoniae* in Ghana and other African countries (Kwarteng et al. 2017; Nuoh et al. 2016; Scarborough et al. 2007). This suggest that if controls and prevention efforts need to be improved such as mass vaccination campaigns to target predominant strains like *Neisseria meningitidis, W135*, and *Streptococcus pneumonia St*.*1. Haemophilus influenzae* (3.3%) with 60.7% of the confirmed strains being *Haemophilus influenzae B*. These finding are in congruent with studies conducted by Muller (2014) which denoted that after the introduction of a conjugated Hib vaccine in 1980, the incidence of Hib infections has decreased by 90%. Currently Hib is responsible for 6.7% of all bacterial meningitis cases [23].

Diagnosis and identification of the isolates causing the bacterial meningitis in the cerebrospinal fluid and the early initiation of the appropriate treatment is the most critical stage in the management of the disease. It is therefore critical to get the best results in order to detect and manage confirmed cases across the country. The Rt-PCR remains the gold standard in Ghana but supplemented by CSF culture for immediate management of cases at the peripheral levels. Studies have shown that the latex agglutination test has a very low sensitivity especially in the patients who have received antibiotic treatment before the lumbar puncture and limits the use of his method [11]. Delays in the diagnosis and treatment are avoided through the routine use of CSF culture methods in the patients under the suspicion of bacterial meningitis [24]. The study of the comparison of the culture to the gold standard of Rt-PCR method revealed that there is a statistically significant (z = - 22.596; p<0.05) difference between the CSF culture results and Rt-PCR methods chained out. In comparison, there was a positive agreement of 11.1% whiles positive disagreement of Rt-PCR was 23.3%. Positive disagreement for CSF culture was 0.2% in the overall samples tested.

Findings from the results showed that the CSF culture results were not in accordance with those of Rt-PCR. Although some performance characteristics were in good correlation with Rt-PCR results such us specificity (98.2%) and positive predictive value (90.5%). Meanwhile, the sensitivity (31.8%) and negative predictive value (72.6%) of CSF culture were low, which means that majority of the CSF culture results were false negative. The findings are favourably similar with the results of other studies who found specificity of CSF culture not to reflect the true percentage, because the presence of fastidious bacteria, delay in CSF culturing, or consumption of antibiotics before lumbar puncture, could result in altering the outcomes [25]–[27]. These outcomes can be attributed to the procedures undertaken before lumbar puncture is performed. Literature states that Rt-PCR is more accurate and reliable, especially among patients who have used antibiotics before lumbar puncture (Ceyhan et al., 2008; Saravolatz et al., 2003). Although some performance characteristics were in good standing by culture results such as the level of agreements of CSF culture with Rt-PCR results, the difference was clearly ascertained by the signed rank test.

## Data Availability

Data collected for this study was obtained from the Tamale Public Health and Reference Laboratory. The data is not open to the public and hence permission is required to have access to it.

## Ethics Approval Statement

Approval was given from the Committee on Human Research, Publication and Ethics with Ethics Number, CHRPE/AP/469/20 from the Kwame Nkrumah University of Science and Technology.

## Acknowledgement

The authors wish to thank the leadership of Ghana Health Service, Public Health Division and the Disease Surveillance Unit for their support and guidance. We also want to thank the leadership of the ZPHL add the rest for the immense support given to us through the period of data collection and processing.

## Authors’ contributions

The authors of the research contributed in diverse areas with specific roles and responsible as denoted;

^1*^Enoch Weikem Weyori, contributed in document management and write up

^2^Abass Abdul-Karim contributed in document review

^3^John Bertson Eleeza contributed in document review

^4^Nkrumah Bernard, contributed in document review

^5^Braimah Baba Abubakari contributed in document review

^6^Hilarius Asiwome Kosi Abiwu contributed in document review

^7^Kuugbee Dogkotenge Eugene contributed in document review

^8^Adadow Yidaana contributed in document review

^9^Benjamin Nuertey contributed in literature review

^10^Adatsi Richard Kojo contributed in laboratory testing and results management

^11^Stebleson Azure contributed in laboratory testing and results management

^12^Sylvester Mensah contributed in laboratory testing and results management

^13^Etowi Boye Yakubu contributed in data management and analysis

^14^Valentine Koyiri Cheba contributed in laboratory testing and sample management

^15^Alentis Aba-ifaa Dombadoh contributed in laboratory testing and sample management

## Data Sharing Statement

Even though ethics approval was obtained from CHRPE and GHS, the data for this study will not be shared due to the fact that the data obtained was a retrospective data of which no form of consent was sort from the patients or participants who sort for medical care and treatment from the respective facilities across the zone.

